# Estimating the Per-Application Effectiveness of Chlorhexidine Gluconate and Mupirocin in Methicillin-resistant *Staphylococcus aureus* Decolonization in Intensive Care Units

**DOI:** 10.1101/19012732

**Authors:** Eric T. Lofgren, Matthew Mietchen, Christopher Short, Kristen V. Dicks, Rebekah Moehring, Deverick Anderson, for the CDC MIND-Healthcare Program

**Affiliations:** Paul G. Allen School for Global Animal Health, Washington State University, Pullman, WA; Duke Center for Antimicrobial Stewardship and Infection Prevention, Durham, NC

## Abstract

**Introduction:** Chlorhexidine gluconate and mupirocin are widely used to decolonize patients with methicillin-resistant *Staphylococcus aureus* (MRSA) and reduce risks of infection in hospitalized populations. The probability that a treated patient would be decolonized, which we term per-application effectiveness, is difficult to directly measure. Quantifying the efficacy of per-application effectiveness of CHG and mupirocin is important for studies evaluating alternative decolonization strategies or schedules as well as identifying whether there is room for improved decolonizing agents.

**Methods:** Using a stochastic compartmental model of an intensive care unit (ICU), the per-application effectiveness of chlorhexidine and mupirocin were estimated using approximate Bayesian computation. Extended sensitivity analysis examined the potential impact of a latent period between MRSA colonization and detection, the timing of decolonization administration, and parameter uncertainty.

**Results:** The estimated per-application effectiveness of chlorhexidine was 0.15 (95% Credible Interval: 0.01, 0.42), while the estimated effectiveness of mupirocin was is 0.15 (95% CI: 0.01, 0.54). A lag in colonization detection markedly reduced both estimates, which were particularly sensitive to the value to the modeled contact rate between nurses and patients. Gaps longer than 24-hours in the administration of decolonizing agents still resulted in substantial reduction of within-ICU MRSA transmission.

**Discussion:** The per-application effectiveness estimates for chlorhexidine and mupirocin suggest there is room for substantial improvement in anti-MRSA disinfectants, either in the compounds themselves, or in their delivery mechanism. Despite these estimates, these agents are robust to delays in administration, which may help in alleviating concerns over patient comfort or toxicity.

## Introduction

Despite recent progress in reducing the incidence of methicillin-resistant *Staphylococcus aureus* (MRSA) in hospitals^1^, it remains a targeted pathogen for infection prevention and public health efforts. One intervention with increasingly widespread use is decolonization of patients with MRSA using chlorhexidine gluconate (CHG) baths for the skin and mupirocin for the nares. While these interventions have shown to be effective in a number of randomized controlled trials^2^, the results from some community-based studies have been more equivocal^3^. There are several possible explanations for this discrepancy. The results from the randomized trials may not generalize well to settings with lower MRSA incidence. Similarly, lower-incidence settings may not be sufficiently powered to detect an effect of implementing decolonization programs. Finally, there may be changes in implementation from the trial setting to everyday use that decreases the overall effect of the intervention. Thus, infection prevention programs considering these strategies in lower incidence settings must justify the cost of the decolonization products and implementation effort in their hospitals. A better understanding of effectiveness on the per patient application level may help weigh the implementation costs of these interventions.

Evaluating these discrepancies requires a mechanistic understanding of the effectiveness of decolonization – that is, what is the probability, if a patient is treated with a decolonization agent, that they are indeed decolonized? This estimate is essential for a number of potential uses: cost-effectiveness studies, quantifying the impact of a decolonization protocol in conjunction with other interventions, or studying the future impact of changes in effectiveness, due to new technology, emerging resistance to decolonizing agents, or other factors. Obtaining such an estimate empirically, especially in a community setting, would be difficult, requiring intensive and repeated sampling of patients with already complicated clinical cases. Rather than directly measuring the probability of successful decolonization, a mathematical modeling approach can define what probability best supports the results seen in the clinical trials – and with what degree of certainty.

The aim of this mathematical modeling study was to estimate the per-application effectiveness of both CHG bathing and CHG bathing in conjunction with mupirocin decolonization of the nares.

## Methods

### MRSA Transmission Model

We adapted a previously published stochastic compartmental model ^4,5^ of transmission of MRSA through an ICU. The model included compartments for patient colonization status and the presence or absence of contamination on the hands or clothing of healthcare workers (HCWs). Patients were modeled as being either presently uncolonized (P_U_) or colonized (P_C_). were modeled as being either presently uncolonized (P_U_) or colonized (P_C_), while HCWs were modeled as being either uncontaminated (S_U_) or contaminated (S_C_). The model assumed that transmission occurred when a contaminated HCW came into contact with an uncolonized patient, and contamination occurred when an uncontaminated HCW came into contact with a colonized patient (Figure 1). As there is considerable evidence that MRSA can be spread via surface-contamination as well as direct contact^6^, we modeled contact between a patient and an HCW as a direct care task^7,8^ involving either interaction with a patient or their immediate surroundings.

**Figure 1.**
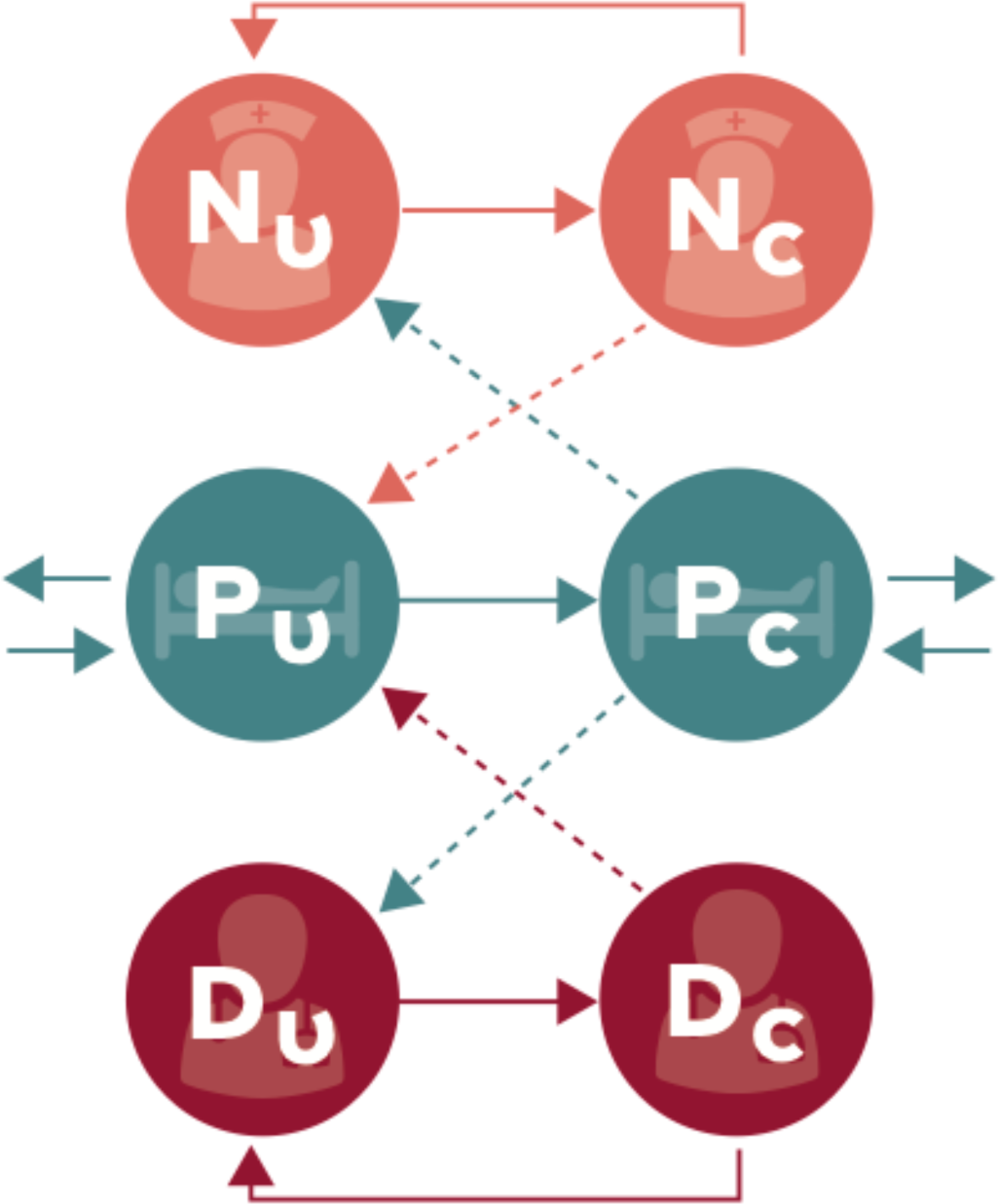
Schematic representation of the compartmental flow of a mathematical model of methicillin-resistant *Staphylococcus aureus* (MRSA) acquisition and CHG/mupirocin decolonization. Solid arrows indicate possible transition states, while dashed arrows indicate potential routes of MRSA contamination or colonization. Nurses and doctors are classified as uncontaminated (NU or DU) and contaminated (NC and DC), while patients are classified as uncolonized (PU) or colonized (PC). Figure by Eric Lofgren is licensed under CC BY 4.0.

We simulated an 18-bed closed ICU assumed to be at maximum capacity, with a 1:3 nurse:patient ratio and a single dedicated intensivist. Because an intensive care unit is a highly structured population, this model relaxes the random mixing assumption used in many compartmental models, instead sub-dividing the patient population such that each patient is cared for by a single nurse and that nurse exclusively cares for three patients. While this is a simplification of the structure of an ICU, previous work has shown it to be a more conservative assumption when compared to random mixing^4^. The intensivist was assumed to treat all patients (Figure 2).

**Figure 2.**
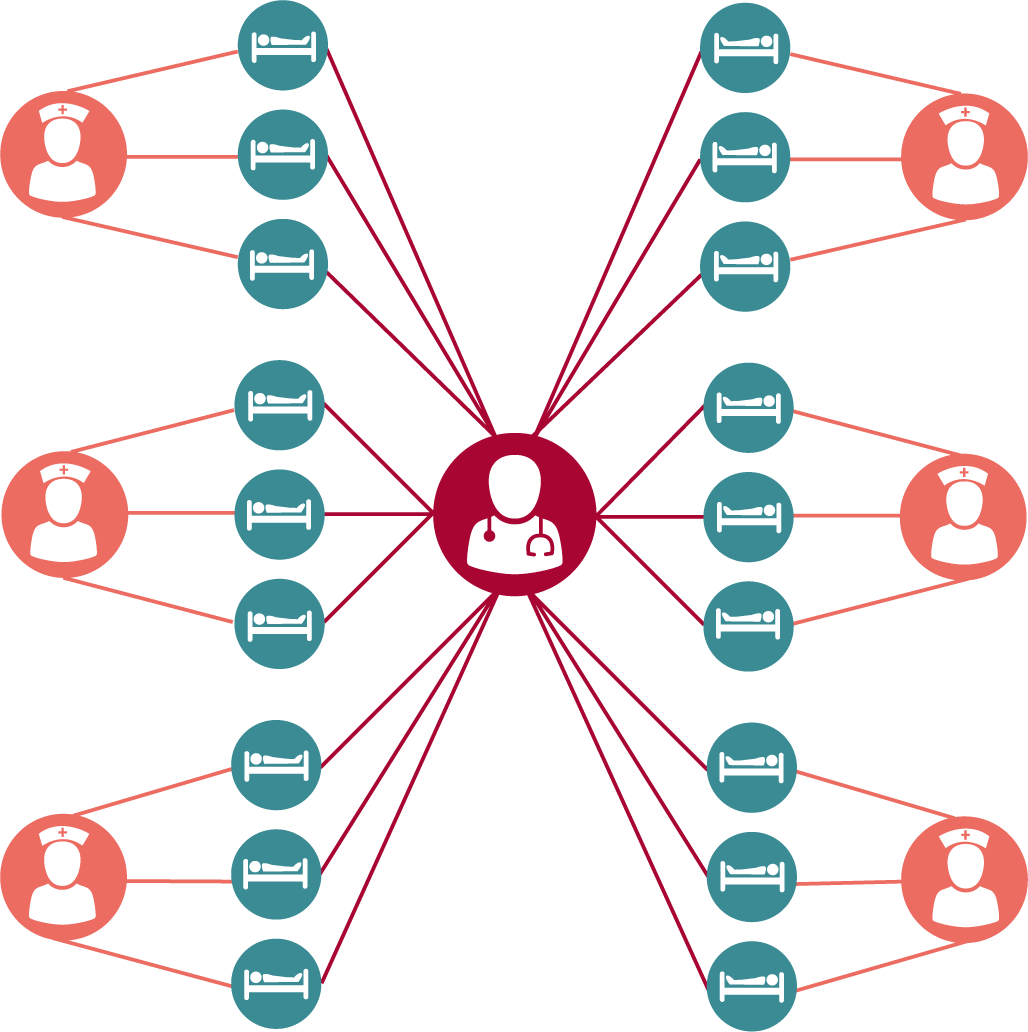
Schematic representation of a structured intensive care unit population to model methicillin-resistant *Staphylococcus aureus* (MRSA) acquisition and CHG/mupirocin decolonization. Patients (blue) are treated by a single assigned nurse (orange). A single intensivist (red) randomly treats all patients. Figure by Eric Lofgren is licensed under CC BY 4.0.

This model makes several simplifying assumptions intended to largely mimic the environment of a hospital with no major outstanding failings in their infection control program. Patients were assumed to be homogeneous in their risk of MRSA acquisition, and the contact frequency between HCWs and patients, while non-random, was uniform (i.e. there are no particularly difficult or contact-intensive patients). Patients were assumed to not interact with each other directly and to be assigned to single-occupancy rooms. Hospitals were assumed to follow standard contact precaution guidelines set forward by the CDC and to detect MRSA colonization with perfect accuracy. Finally, all HCWs were assumed to wash their hands after each direct care task and to change their gloves and/or gowns at a rate equal to when entering and exiting the patient room. These assumptions are intended to largely mimic the environment of a hospital with no major outstanding failings in their infection control program.

### Parameterization and Model Calibration

The model largely used parameter values from a previously published model^4,5^. The values of each parameter in the model, and the source they were drawn from, are detailed in Table 1. The stochastic reaction equations that govern the model and code necessary to run the simulations are available at https://github.com/epimodels/chg_effectiveness. The transition terms are provided in Supplemental Appendix A. Where possible, parameters were drawn from studies from large academic medical centers similar to the ones conducting large RCTs on decolonization protocols.

**Table 1.**
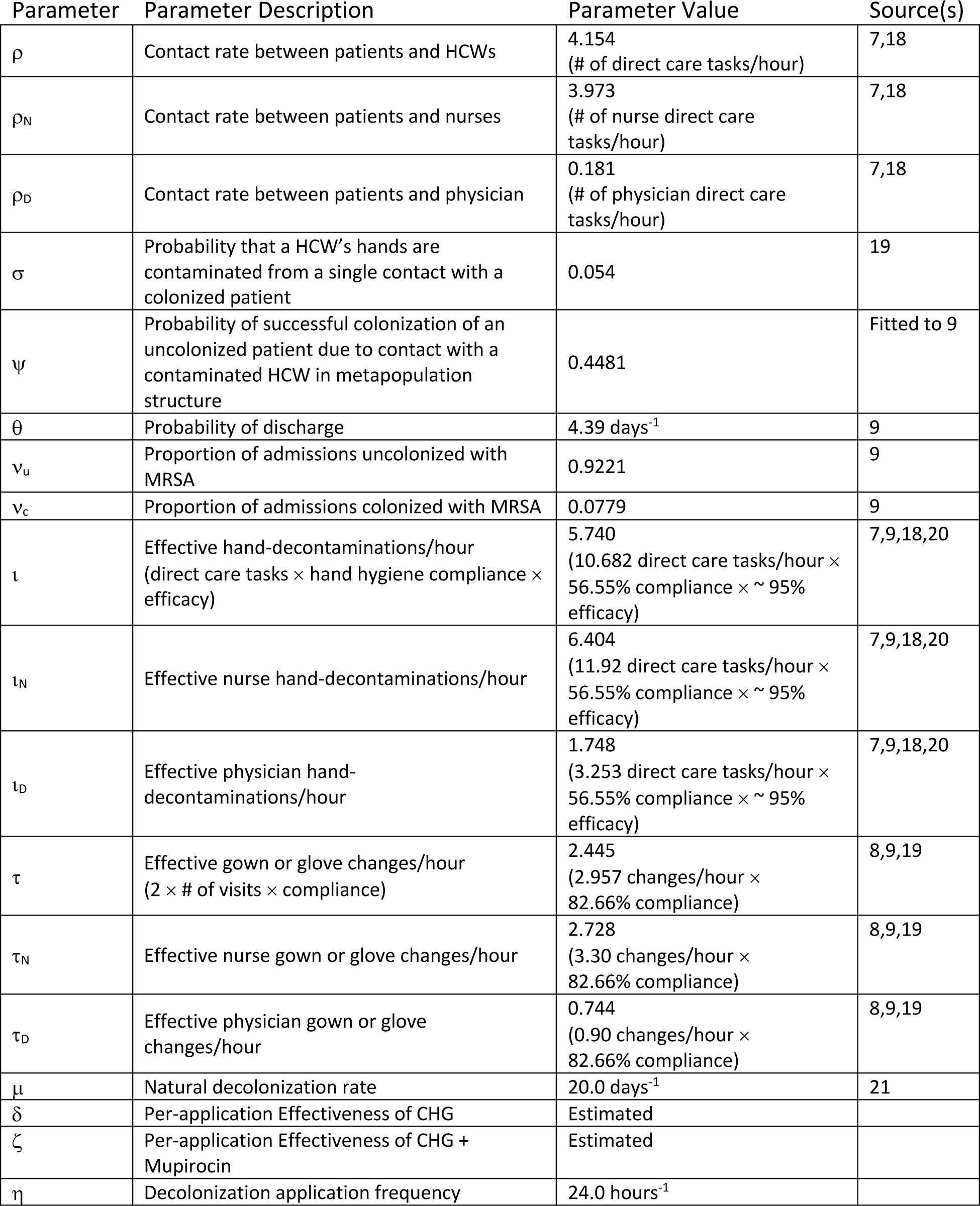

| Parameter | Parameter Description | Parameter Value | Source(s) |
| --- | --- | --- | --- |
| $\rho$ | Contact rate between patients and HCWs | 4.154<br>(# of direct care tasks/hour) | 7,18 |
| $\rho_N$ | Contact rate between patients and nurses | 3.973<br>(# of nurse direct care tasks/hour) | 7,18 |
| $\rho_D$ | Contact rate between patients and physician | 0.181<br>(# of physician direct care tasks/hour) | 7,18 |
| $\sigma$ | Probability that a HCW's hands are contaminated from a single contact with a colonized patient | 0.054 | 19 |
| $\psi$ | Probability of successful colonization of an uncolonized patient due to contact with a contaminated HCW in metapopulation structure | 0.4481 | Fitted to 9 |
| $\theta$ | Probability of discharge | 4.39 days<sup>-1</sup> | 9 |
| $v_u$ | Proportion of admissions uncolonized with MRSA | 0.9221 | 9 |
| $v_c$ | Proportion of admissions colonized with MRSA | 0.0779 | 9 |
| $\iota$ | Effective hand-decontaminations/hour<br>(direct care tasks × hand hygiene compliance × efficacy) | 5.740<br>(10.682 direct care tasks/hour × 56.55% compliance × ~ 95% efficacy) | 7,9,18,20 |
| $\iota_N$ | Effective nurse hand-decontaminations/hour | 6.404<br>(11.92 direct care tasks/hour × 56.55% compliance × ~ 95% efficacy) | 7,9,18,20 |
| $\iota_D$ | Effective physician hand-decontaminations/hour | 1.748<br>(3.253 direct care tasks/hour × 56.55% compliance × ~ 95% efficacy) | 7,9,18,20 |
| $\tau$ | Effective gown or glove changes/hour<br>(2 × # of visits × compliance) | 2.445<br>(2.957 changes/hour × 82.66% compliance) | 8,9,19 |
| $\tau_N$ | Effective nurse gown or glove changes/hour | 2.728<br>(3.30 changes/hour × 82.66% compliance) | 8,9,19 |
| $\tau_D$ | Effective physician gown or glove changes/hour | 0.744<br>(0.90 changes/hour × 82.66% compliance) | 8,9,19 |
| $\mu$ | Natural decolonization rate | 20.0 days<sup>-1</sup> | 21 |
| $\delta$ | Per-application Effectiveness of CHG | Estimated | |
| $\zeta$ | Per-application Effectiveness of CHG + Mupirocin | Estimated | |
| $\eta$ | Decolonization application frequency | 24.0 hours<sup>-1</sup> | |

### Decolonization Intervention Efficacy Estimation

In order to estimate the per-application effectiveness of a CHG and/or a CHG-Mupirocin combination intervention, we used a three-step fitting procedure: baseline, intervention 1 (CHG baths), and intervention 2 (CHG baths plus nasal mupirocin). First, a baseline model of a pre-intervention intensive care unit was fit using a single free parameter, ψ, that governs the probability an uncolonized patient becomes colonized after contact with contaminated healthcare worker. This parameter was tuned such that the model had an average incidence of 5.89 MRSA acquisitions per 1000 patient-days^5,9^. Second, we introduced a new parameter, d, to represent CHG-based decolonization, when a patient moved patients from a colonized status to an uncolonized status (Intervention 1). This parameter was assumed to result in a 0.748 incidence rate ratio compared to the baseline model, in line with a meta-analysis of CHG-only studies by Kim *et al*^2^. Third, a second parameter which moved patients from colonized to uncolonized, ζ, was fit to represent the addition of nasal decolonization with mupirocin accompanying a CHG bathing protocol (Intervention 2), resulting in a combined incident rate ratio of 0.578. This formulation assumes that the effects of CHG and mupirocin are additive and that there is no synergistic effect between them.

Approximate Bayesian Computation (ABC) was used to fit these parameters. Details on ABC for model fitting may be found elsewhere^10,11^. Briefly, ABC is a computational technique that draws a candidate value from a prior distribution, simulates the model using that value, and accepts the candidate value if the simulated result is within an error band around a given summary statistic. In this case, we fit the model to incidence rates corresponding to the baseline and the two simulated interventions. The distribution of these accepted values is an approximation of a Bayesian posterior. In this study, all parameters were fit using 1,000,000 parameter draws from a uniform prior distribution bounded by 0.0 and 1.0. Candidate parameters were accepted with an error term, ε = 0.05, indicating that the simulated incidence rates had to be within ± 5% of the target incidence rate on the log scale.

### Sensitivity Analysis

Three separate sensitivity analyses were conducted. The first varied the frequency with which decolonization was applied, comparing a baseline of no decolonization to applications of CHG and mupirocin every 24, 48, 72, 96, and 120 hours to see if a substantial portion of the modeled efficacy is dependent on the typical schedule of a daily CHG bath.

The second was a global sensitivity analysis, simultaneously allowing each parameter to vary uniformly ± 50% of its original value. For each parameter draw, 200 model runs were performed and the joint efficacy of CHG and mupirocin (as a single parameter) was re-estimated. This process was repeated 5,000 times, and linear regression was used to estimate the relative impact of a single percentage change in each parameter value on the estimated efficacy.

Finally, we conducted a structural sensitivity analysis examining the impact of assuming – as our model did – that there is no latent period in MRSA colonization, wherein a patient is colonized at sub-detectable levels. We added a latent period to our model, wherein patients transitioned from P_s_ to a new compartment, P_E_ – representing latent colonization–before finally transitioning to the P_c_ colonized state. The rate of transition from P_E_ to P_c_ varied randomly from one to four days12. Patients in P_E_ were assumed not to shed sufficient amounts of MRSA to contaminate healthcare workers. Effectively, this creates a small pool of patients who, despite being decolonized due to treatment, are not recognized as such, as their MRSA acquisition has not yet been detected. The per-application efficacy of CHG and mupirocin were then re-estimated using the same procedures as the main model.

## Results

### Per-Application Efficacy of CHG Bathing and CHG-Mupirocin Combinations

The estimated per-application efficacy of CHG bathing to induce colonization rates similar to those seen in Kim *et al*. was 0.15 (95% Credible Interval (CI): 0.01, 0.42), meaning a little under a sixth of all applications of CHG are expected to result in effective decolonization of the patient. Mupirocin had an estimated per-application effectiveness of 0.15 (95% CI: 0.01, 0.54). The posterior densities of the efficacy estimates are shown in Figure 3. The addition of a 1 to 4-day latent period in between the transmission event and recognized MRSA colonization reduced both efficacy estimates. In this case, CHG and Mupirocin had estimated per-application efficacies of 0.11 (95% CI: 0.01, 0.30) and 0.10 (0.004, 0.34) respectively (Figure 3).

**Figure 3.**
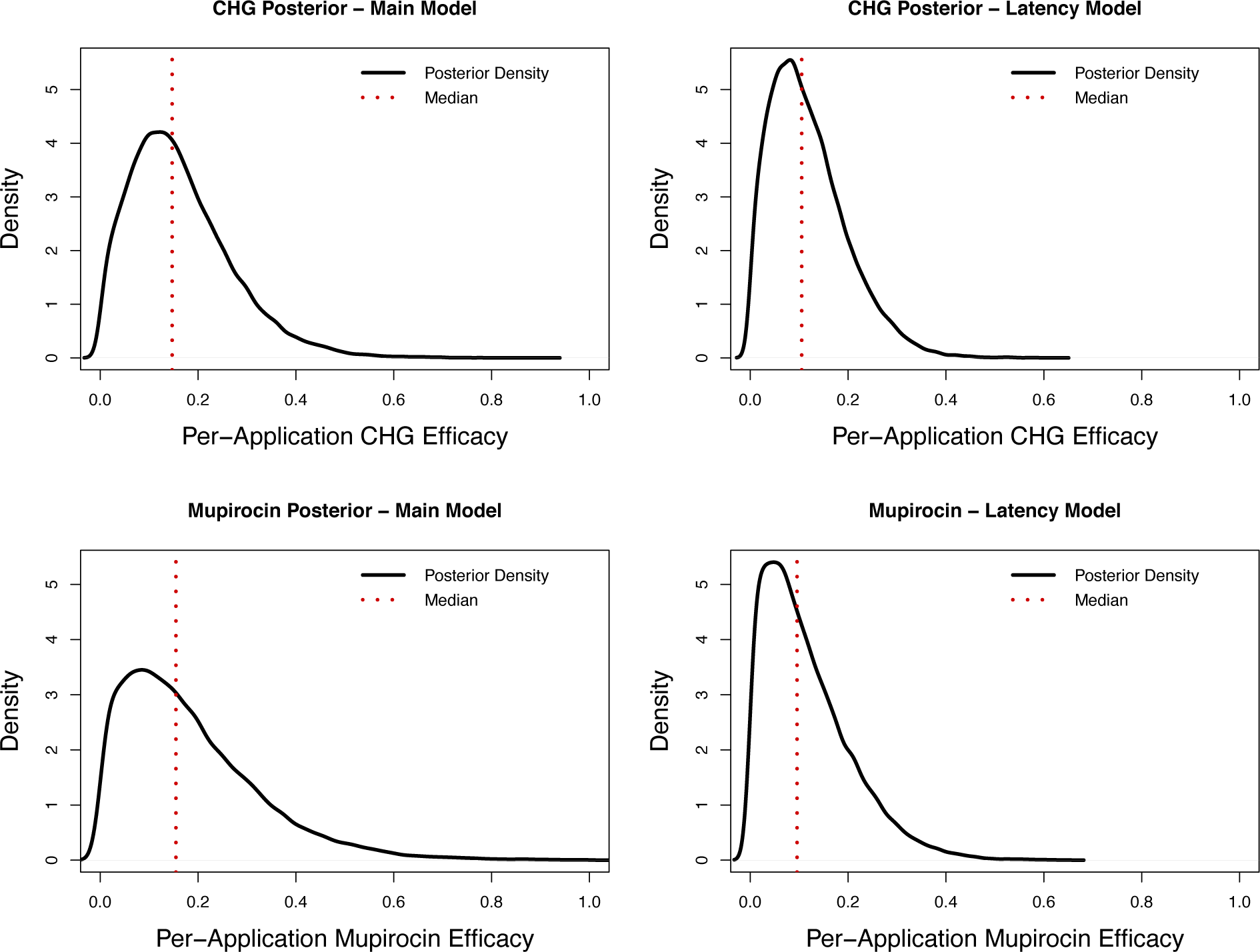
Approximate Bayesian Posterior Estimates for Per-Application Chlorhexidine Gluconate (delta) and Mupirocin (zeta) Effectiveness. Each panel shows the density of accepted values (dark line) and the median of this density (dotted line). Densities were estimated using a normal kernel. Left-hand panels show the estimates assuming acquisition is instantly detected, while the right-hand panels show the estimates assuming there is a one to four-day latent period where a patient may be colonized (and decolonized) but their acquisition is not yet detected.

### Model Sensitivity to Variation in Timing and Parameter Uncertainty

Despite this relatively modest per-application efficacy estimate, the results of the timing sensitivity analysis showed that substantial decreases in MRSA acquisitions can be achieved at much less frequent bathing intervals. Compared to a mean of 1.23 acquisitions per 1,000 patient-days in the control scenarios, a bathing protocol administering CHG and mupirocin every 120 hours (5 days) resulted in a mean acquisition rate of 1.03 acquisitions per 1,000 patient days, a 16.3% decrease (p > 0.001) (Figure 4).

**Figure 4.**
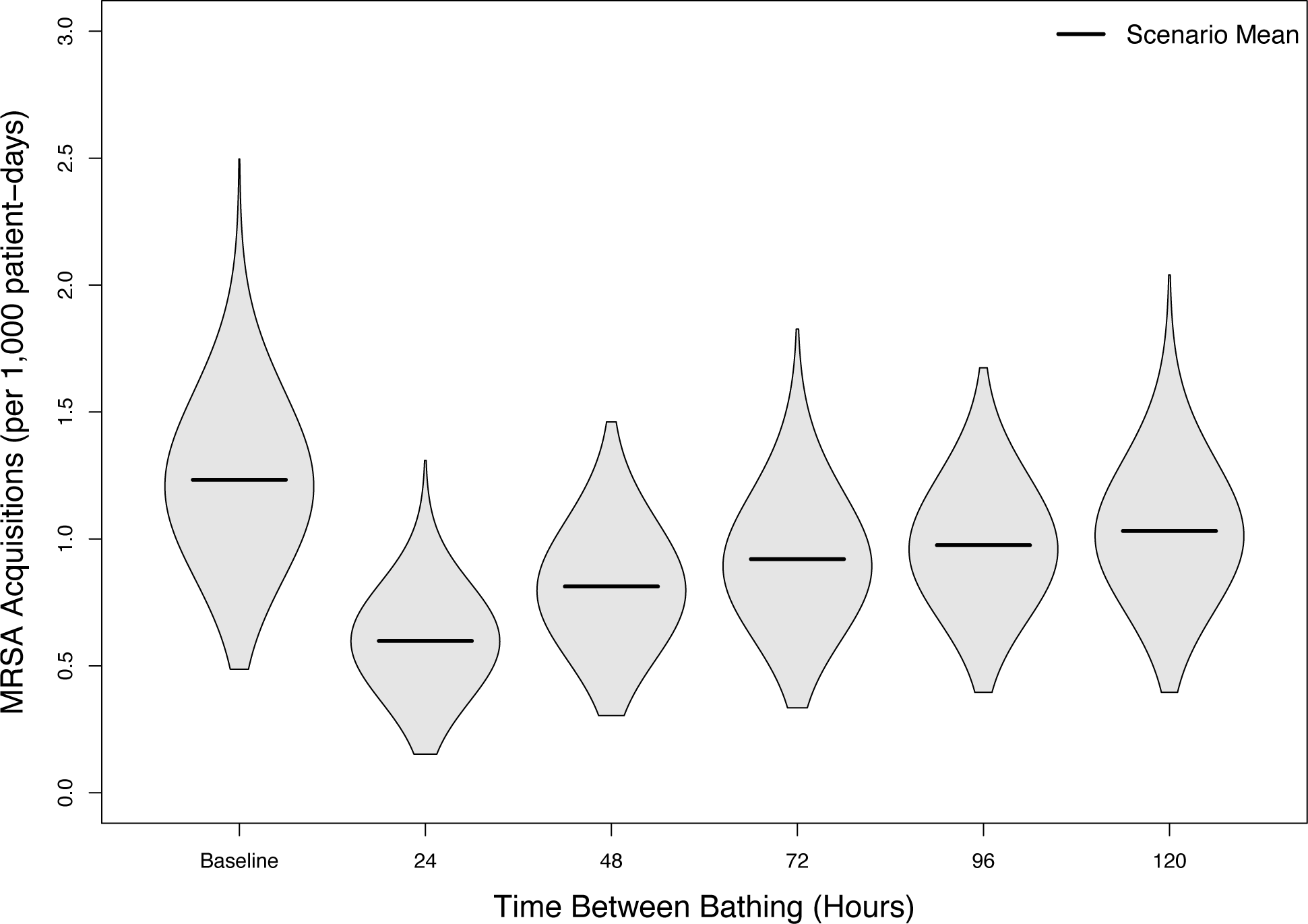
Violin plot of the Sensitivity of Decolonization Protocols to Changes in Timing. Each ‘violin’ shows a smoothed kernel-density estimate of 1,000 runs of the model with a given timing for the administration of decolonizing baths, in acquisitions per 1,000 patient-days.Solid, black, horizontal bars indicate the mean estimate for each scenario.

The model’s results were most sensitive to the value of *ρ*_*N*_, the contact rate between nurses and patients. A 1% increase in the value of this parameter corresponded to a 0.73% increase in the estimated combined efficacy of CHG and mupirocin (95% CI: 0.71, 0.75). Other sensitive parameters included y, the probability of colonization given contact between a contaminated HCW and a patient (0.43% (95% CI: 0.41, 0.44) and n, the proportion of admissions colonized with MRSA (0.37% (95% CI: 0.35, 0.39). The sensitivity estimates for all varied parameters is shown in Figure 5.

**Figure 5.**
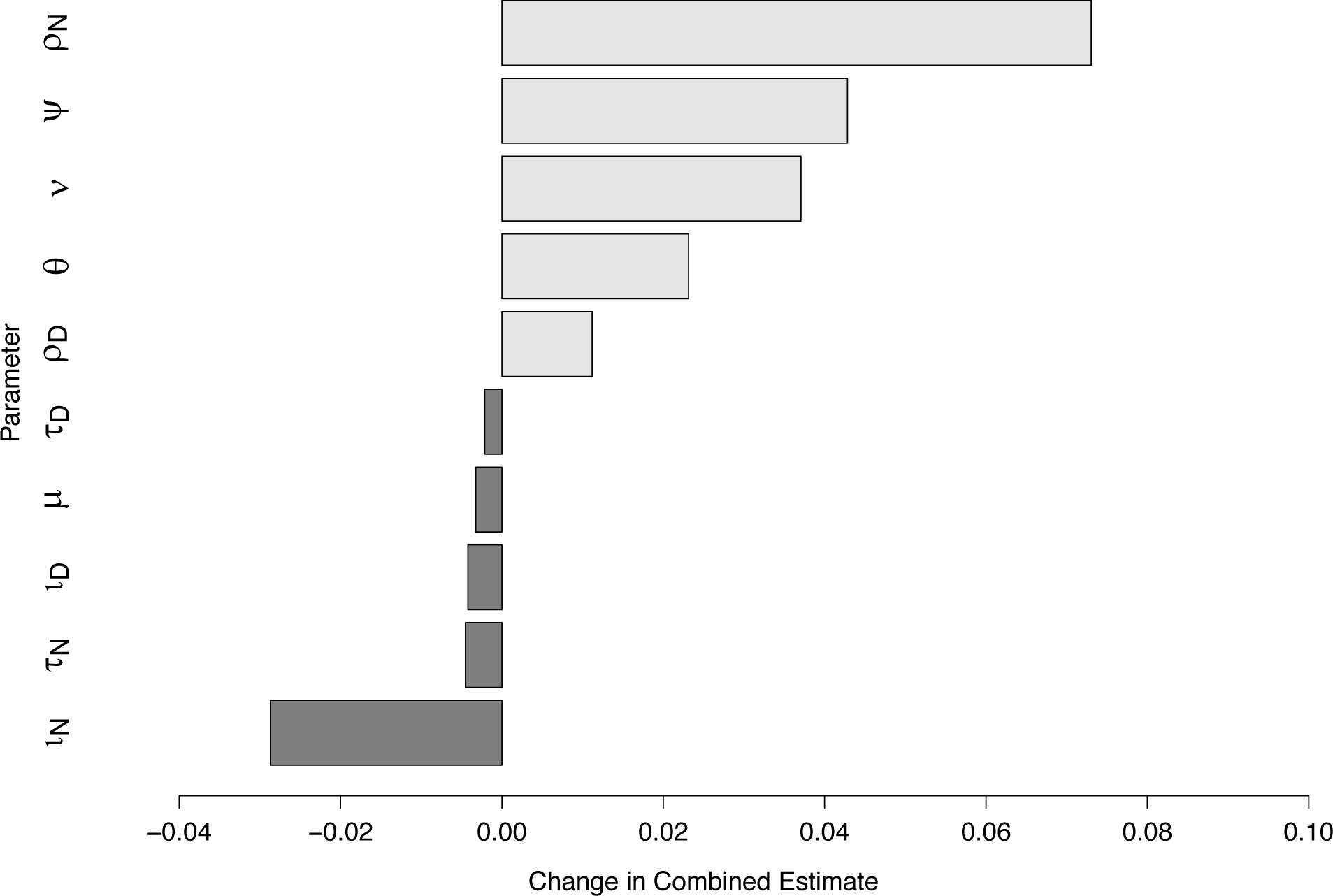
Global sensitivity of a mathematical model of methicillin-resistant *Staphylococcus aureus* (MRSA) acquisition and CHG/mupirocin decolonization. Horizontal bars represent the change in the estimated effectiveness of CHG/mupirocin decolonization per one-percent change in the value of a specific parameter, with light bars indicating increased estimated effectiveness and dark bars indicating decreased estimated effectiveness.

## Discussion

Using a mathematical model to translate from population-level effect estimates to a per-application effectiveness estimate, this study suggests that on a per-application basis both CHG and mupirocin are at best mildly effective at decolonizing patients with MRSA. Under ideal circumstances, the combination of the two compounds was estimated to be effective 30% of the time. Under more realistic circumstances where there is some delay and uncertainty between the acquisition and detection of MRSA, either due to biological processes surrounding colonization or laboratory testing, this estimate drops dramatically to 20%.

These results should not be taken as a condemnation of the utility of CHG as an option for reducing the transmission of MRSA within hospitals. Rather, it illustrates that even relatively imperfect interventions may still have impact. Further, it suggests that there may be room for substantial further gains by improving the methods by which we decolonize patients.

Importantly, this model cannot distinguish whether or not the effectiveness of CHG and mupirocin are due to the compounds themselves or the way in which they are applied. This means that, even in the absence of novel compounds, improvements to the methods of applying CHG and mupirocin may reap considerable benefits^13,14^.

The results of the timing-focused sensitivity analysis show that considerable deviations from an intensive 24-hour decolonization schedule can still result in substantial reductions in the unit-level MRSA acquisition rate. Most healthcare-associated pathogens have relatively low transmissibility^15–17^, and as such any reduction in the colonization pressure within an ICU, even a modest one, can interrupt delicate transmission chains. This study suggests that deviations from a daily CHG bathing schedule due to concerns over toxicity in pediatric populations, patient-reported skin irritation, or other practical demands are still potentially useful interventions. These results are also potentially useful for future studies, allowing facilities to estimate the expected impact in their specific settings, allowing for the critical evaluation of existing studies, and providing clear estimates that can be used to estimate the impact of resistance to CHG, mupirocin, or both.

This study is not without limitations. Broadly, it assumes that the ICU represented in the model, which is meant to represent the type of academic medical center where large-scale intervention trials are most often conducted, is a reasonable representation of the environment in which the studies were actually conducted. The parameter sensitivity analysis shows that the model is most sensitive to errors in the contact rate between nurses and patients. Further, like the meta-analysis by Kim *et al*. that was used to calibrate the model and estimate the per-application effectiveness of CHG and mupirocin, this study assumes that the studies in question, a mix of randomized trials and interrupted time-series studies, were capable of estimating the population-level impact of decolonization without bias. Additionally, this model assumes there is no cumulative benefits to repeated bathing – each application is treated as a separate and independent event.

Despite these limitations, this study represents an innovative use of mathematical modeling to estimate the effectiveness of a hospital epidemiology intervention using summary statistics to estimate an individual-level effect. In particular, estimating the per-application efficacy of these compounds would be difficult, if not impossible, to directly measure in a working healthcare setting. It shows that there are still substantial prospects for improved decolonization interventions to further reduce MRSA rates in the ICU, and that there is room for deviation from intensive daily protocols in response to patient or clinician needs without overly jeopardizing their impact.

## Data Availability

Simulation code, data and analysis code is available on a public Github repository.

https://github.com/epimodels/chg_effectiveness

## Acknowledgements

This work was supported by the CDC Cooperative Agreement RFA-CK-17-001-Modeling Infectious Diseases in Healthcare Program (MInD-Healthcare). We thank the members of the MInD-Healthcare network for their advice and input.

